# The impact of COVID-19 on pregnant and recently pregnant women in Malawi: A national facility-based cohort

**DOI:** 10.1101/2022.03.15.22272348

**Authors:** Chikondi Chapuma, Leonard Mndala, Luis Gadama, Fannie Kachale, Andrew Likaka, Rosemary Bilesi, Malangizo Mbewe, Bertha Maseko, Chifundo Ndamala, Regina Makuluni, Annie Kuyeri, Laura Munthali, Deborah A. Phiri, Clemens Masesa, Marc Y.R Henrion, Moses Kumwenda, David Lissauer

**Author notes:** shared first authorship (Chikondi Chapuma, Leonard Mndala).

## Abstract

**Objective:** To describe the demographic characteristics, clinical manifestations, and clinical outcomes of hospitalised pregnant and recently pregnant women with COVID-19 in Malawi, a low-income country in Sub-Saharan Africa. This study responds to a critical gap in the global COVID-19 data.

**Methods:** A national surveillance platform was established in Malawi by the Ministry of Health to record the impact of COVID-19 on pregnant and recently pregnant women and provide real-time data for decision making. We report this facility-based cohort that includes all pregnant and recently pregnant hospitalised women in Malawi suspected of having COVID-19 between 2nd June 2020 and 1st December 2021.

**Results:** 398 women were admitted to hospital with suspected COVID-19 based on presenting symptoms and were tested; 246 (62%) were confirmed to have COVID-19. In women with COVID-19, the mean age was 27 ± 7 years.

The most common presenting symptoms were cough (74%), breathlessness (45%), Fever (42%), headache (17%), and joint pain (10%). 53% of the women had COVID-19 symptoms severe enough to warrant admission.

31% (76/246) of women admitted with COVID-19 suffered a severe maternal outcome, 47/246 (19%) died, and 29/246 (12%) had a near-miss event. 9/111 (8%) of recorded births were stillbirths, and 12/101 (12%) of the live births resulted in early neonatal death.

**Conclusion:** A national electronic platform providing real-time information on the characteristics and outcomes of pregnant and recently pregnant women with COVID-19 admitted to Malawian government hospitals. These women had much higher rates of adverse outcomes than those suggested in the current global data. These findings may reflect the differences in the severity of disease required for women to present and be admitted to Malawian hospitals, limited access to intensive care and the pandemic’s disruption to the health system.

**SUMMARY BOX:** *What is already known?:*

- In pregnant and recently pregnant women, COVID-19 is associated with increased complications such as admission to an intensive care unit, invasive ventilation, and maternal death.
- In pregnant women with confirmed COVID-19, the current global estimate of all-cause mortality is 0.02%.
- Most countries in Africa rely on paper-based systems to collect key maternal health indicators such as maternal deaths and severe morbidity, which does not enable timely actions.

*What are the new findings?:*

- Maternal mortality and adverse perinatal outcomes are alarmingly high in a cohort of pregnant and recently pregnant women admitted to Malawian healthcare facilities located in a low-income country in Africa.
- A national facility-based maternal surveillance platform can be implemented during a pandemic and provide real-time data to aid policymakers in understanding its impacts on this key population.

*What do the new findings imply?:*

- In low-income countries in Sub-Saharan Africa, pregnant and recently pregnant women with COVID-19 admitted to hospital require enhanced care and a renewed focus on their needs to avert these adverse health outcomes.
- Global and national surveillance systems must specifically record outcomes for pregnant, recently pregnant women and their infants to understand the impact of public health emergencies on these groups, as they may be disproportionately affected and may require special considerations.

## INTRODUCTION

Coronavirus disease 2019 (COVID-19) ‘s impact has been felt globally (1). Pregnant women were anticipated to have more severe disease outcomes when infected with COVID-19 due to maternal physiological and immunological changes of pregnancy (2).

We know that in pregnant and recently pregnant women, COVID-19 is associated with increased rates of complications such as admission to an intensive care unit, invasive ventilation, and maternal death. This has been particularly seen in women of increased maternal age, high body mass index, non-white ethnicity, and any pre-existing maternal comorbidity (including chronic hypertension, diabetes, and pre-eclampsia) (3,4). However, the comprehensive living systematic review summarising such data includes no studies from the African region, amongst the 192 included studies(3,4). Budhram et al., in 2021, described maternal characteristics and pregnancy outcomes of hospitalized pregnant women with SARS-CoV-2 virus infection in South Africa (5). This is in the context of a middle-income country, and lead clinicians of hospitals participated on a voluntary basis risking under-reporting of key variables. In addition, even with increasing numbers of maternal COVID-19 cohorts now reported globally (991 on 14/02/2022), none were from low income countries in Africa, and only four appear to be from low-middle income countries (LMICs) in Africa (Kenya, Nigeria, Angola, and Egypt)(6). A multinational cohort including populations from Nigeria and Ghana has also been studied. However, data has not yet been reported to enable the outcomes to be examined at a country level. It was though suggested in this multi-country cohort that no systematic pattern of differences in maternal outcomes were measured between participating countries (7). Therefore, the direct impact of COVID-19 on pregnant and recently pregnant women in Africa, and thus the true global impact, is still not well described (8).

Beyond the direct impact of COVID-19 on pregnant women who become infected, the pandemic also has a serious impact on the safe delivery of pregnancy care, including quality and access to care (9).Hence, pregnant women have been impacted due to the wider health system strain, and social and economic consequences of COVID-19, and of the societal responses to COVID required to minimise spread. For example, early in the pandemic, the national lockdown in Nepal was associated with an increase in the institutional stillbirth rate and neonatal mortality, and a decrease in the quality of care (10). Malawi had registered 62,088 confirmed COVID-19 cases, 2,307 COVID-19 related deaths and 58,846 recoveries as of 10^th^ December 2021 (11). These absolute numbers of reported cases may also reflect limited testing capacity and uptake, as sero-surveillance has demonstrated that 51.1% of the population had serological evidence of prior infection in February,2021, which is now reported to be as high as 80% in the city of Blantyre (southern region) (12). The pandemic has challenged the country’s healthcare system, resulting in a disruption in the delivery of quality healthcare (13). For instance, an analysis of TB notification data during COVID-19 for Blantyre, Malawi, reported a 36% reduction in TB notifications immediately after the first cases of COVID-19 were reported, with women and girls being more impacted than men and boys (13).

Notably, Malawi’s Ministry of Health (MoH) prioritised the response towards the pandemic but this has been met with numerous existing challenges within the healthcare system (14). In response to the global pandemic, and to monitor in real-time the impact of COVID-19 on maternal health in Malawi, in 2020, the Malawian Ministry of Health, in collaboration with the Malawi Liverpool Wellcome Research Programme, implemented an online national maternal surveillance system (MATSurvey platform) for capturing data on the direct and indirect effects of COVID-19 on maternal and neonatal outcomes in 33 healthcare facilities. Data collected through the platform is made available in real-time through an online interface to designated national, regional and district policy makers, managers and clinicians (Appendix I). This national data collection platform creates a unique opportunity to address the COVID-19 data gap related to the impacts on pregnant women in Africa.

This study describes the demographic characteristics, clinical manifestations, and clinical outcomes of hospitalised pregnant and recently pregnant women with COVID-19 in a low-income country (LIC) in sub-Saharan Africa (SSA), Malawi. The findings of this study will contribute to global efforts in understanding and modelling the impact of the COVID-19 pandemic by providing SSA specific data (4).

## METHODS

### Study Design

This study utilised data gathered by the MATSurvey platform, established by the Ministry of Health, with technical and implementation support from the Malawi-Liverpool-Wellcome Research Programme. Data gathered through the platform was fully anonymised and made available to the research team by permission of the Ministry of Health, Malawi and the College of Medicine Research Ethics committee (COMREC, P.11/20/3186).

This is a facility-based cohort study of pregnant and recently pregnant women in Malawi who were suspected and tested for SARS-CoV-2 from the MATSurvey platform from 2nd June 2020 to 1^st^ December 2021 (inclusive of the second and third wave of the COVID-19 Pandemic in Malawi-https://covid19.who.int/region/afro/country/mw) in 33 health facilities. The MATSurvey platform collects data from all central and district government hospitals in Malawi for women with suspected or confirmed COVID-19, this includes maternal and neonatal demographic characteristics, presenting features, test details and clinical outcomes collected at hospital admission and discharge or death.

The MATSurvey platform data is entered for individual women who have suspected or confirmed COVID-19 at hospital admission or if these conditions occur during the patients hospital stay (Figure 1). The patient’s condition is then updated daily until death or discharge using a tablet-based application developed by the Malawi-Liverpool-Wellcome Research Programme. Additional facility level data is collected weekly. All data entry was completed by government employed “safe motherhood coordinator” midwives or their designated deputies. In person training was provided on platform use to site staff from 27^th^ July - 5^th^ August 2020 and additional follow-up training is given as required in person or remotely. Project coordinators designated for each region provided oversight of the platform, working alongside Ministry of Health staff from the Quality Management Directorate and Reproductive Health Directorate. Data cleaning and quality checks are conducted on a weekly basis.

**Figure 1:**
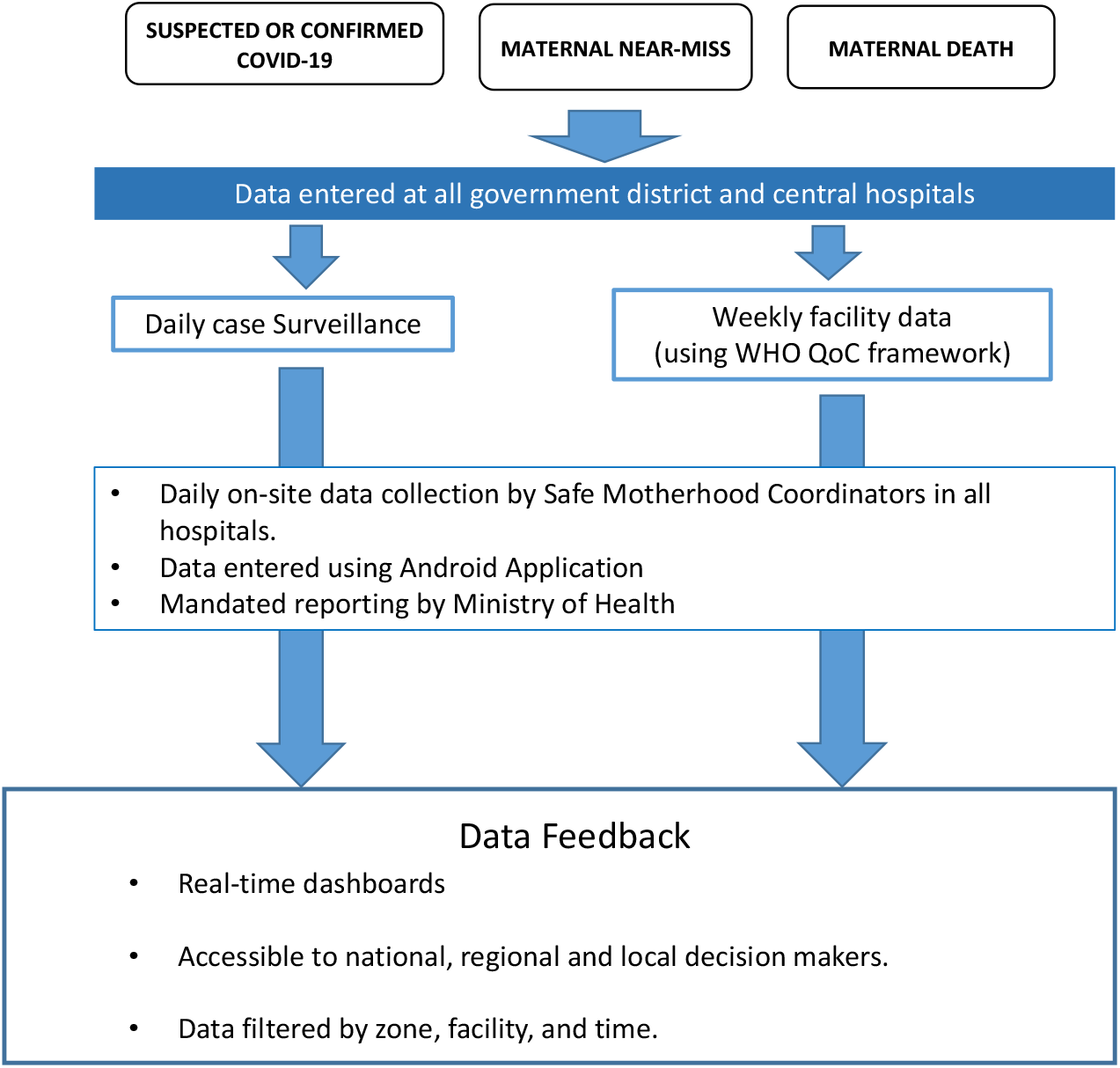
MATSurvey platform Schematic. WHO QoC (Quality of Care) Framework (22)

All data used for this analysis was anonymised to maintain patients’ confidentiality.

### Cohort Selection and Outcomes

A pregnant and a recently pregnant woman was characterised as a COVID-19 suspect based on World Health Organisation’s (WHO) guidance as of May 2020 (15). This required the women to have an acute respiratory illness (fever and a sign of respiratory disease, e.g., cough or shortness of breath) with either travel history or living in the area of community transmission within 14 days or contact with a confirmed or probable case of COVID-19 within 14 days of hospitalization and absence of alternative diagnosis that thoroughly explains the clinical presentation. They also had to be hospitalised at one of the participating facilities (irrespective of whether the hospital admission was COVID-19 related or due to another condition) and be either pregnant or within 6 weeks of the end of pregnancy.

Women were classified as “confirmed COVID-19 cases” if they were confirmed as COVID-19 infected by RT-PCR, gene Xpert testing, or lateral-flow test in a government-approved laboratory. Women were classified as “confirmed COVID negative cases” if, following suspicion of COVID infection, subsequent PCR or gene Xpert or lateral-flow test for SARS-CoV-2 was negative.

### Data Analysis

We performed data cleaning using an open-source relational database management system, the Structured Query Language (MySQL). The data analysis was performed in R version 4.1.1. The demographic characteristics included age, zone/region, and parity. The clinical characteristics included current pregnancy state, pre-existing chronic illnesses, number of antenatal visits, HIV infection status, SARS-CoV-2 related symptoms. The clinical outcomes were severe COVID-19 related outcomes, maternal outcomes (maternal near-miss, maternal death and no adverse outcomes), birth outcomes, and neonatal death.

### Patient and Public Involvement

Overall management of the MATSurvey platform was under the authority of a steering committee which included leadership from directors of the responsible Ministry of Health departments, academic partners and other key in-country health delivery partners such as the Malawi National Blood Transfusion Service. Officials from the Ministry of Health and all districts had access to emerging data through an online dashboard. The Malawi-Liverpool-Wellcome Research Programme, Maternal Health patient and public involvement group are engaged in the dissemination of these findings and wider COVID-19 awareness activities within Malawi.

## RESULTS

This study enrolled patients from 33 healthcare facilities across all regions of Malawi. From 2nd June 2020 to 1^st^ December 2021, a total of 398 pregnant and recently pregnant women met the inclusion criteria of a suspected COVID-19 case. Confirmatory diagnostic testing revealed that 62% (n = 246) were positive for SARS-CoV-2.

### Demographic Characteristics of pregnant and recently pregnant women with COVID-19 infection

The demographic characteristics of women with COVID-19 infection are shown in Table 1. The mean age of women was 27 ± 7 years (mean ± SD), with the majority (30%) aged 20-24 years. Half (n = 124) of the women were hospitalised at facilities in the South-West region of Malawi and 63% were multiparous.

**Table 1:** Demographic Characteristics of pregnant and recently pregnant women with SARS-CoV-2 in Malawi.

| Characteristic | Confirmed COVID-19 (N=246)<br>n (%) |
| --- | --- |
| **Age in years (mean (SD))** | 27 (7) |
| **Age groups** |  |
| <19 | 28 (11) |
| 20-24 | 75 (30) |
| 25-29 | 40 (16) |
| 30-34 | 62 (25) |
| 35-39 | 31 (14) |
| ≥ 40 | 10 (4) |
| **Region** |  |
| Northern | 23 (9) |
| Central East | 26 (11) |
| Central West | 36 (15) |
| South East | 37 (15) |
| South West | 124 (50) |
| **Parity** |  |
| Primigravida | 88 (36) |
| Multigravida | 155 (63) |
| Missing data | 3 (1) |

### Clinical presentation of pregnant and recently pregnant women with COVID-19 infection

The clinical presentation including the history and examination findings of women with COVID-19 disease are shown in Table 2.

**Table 2:**
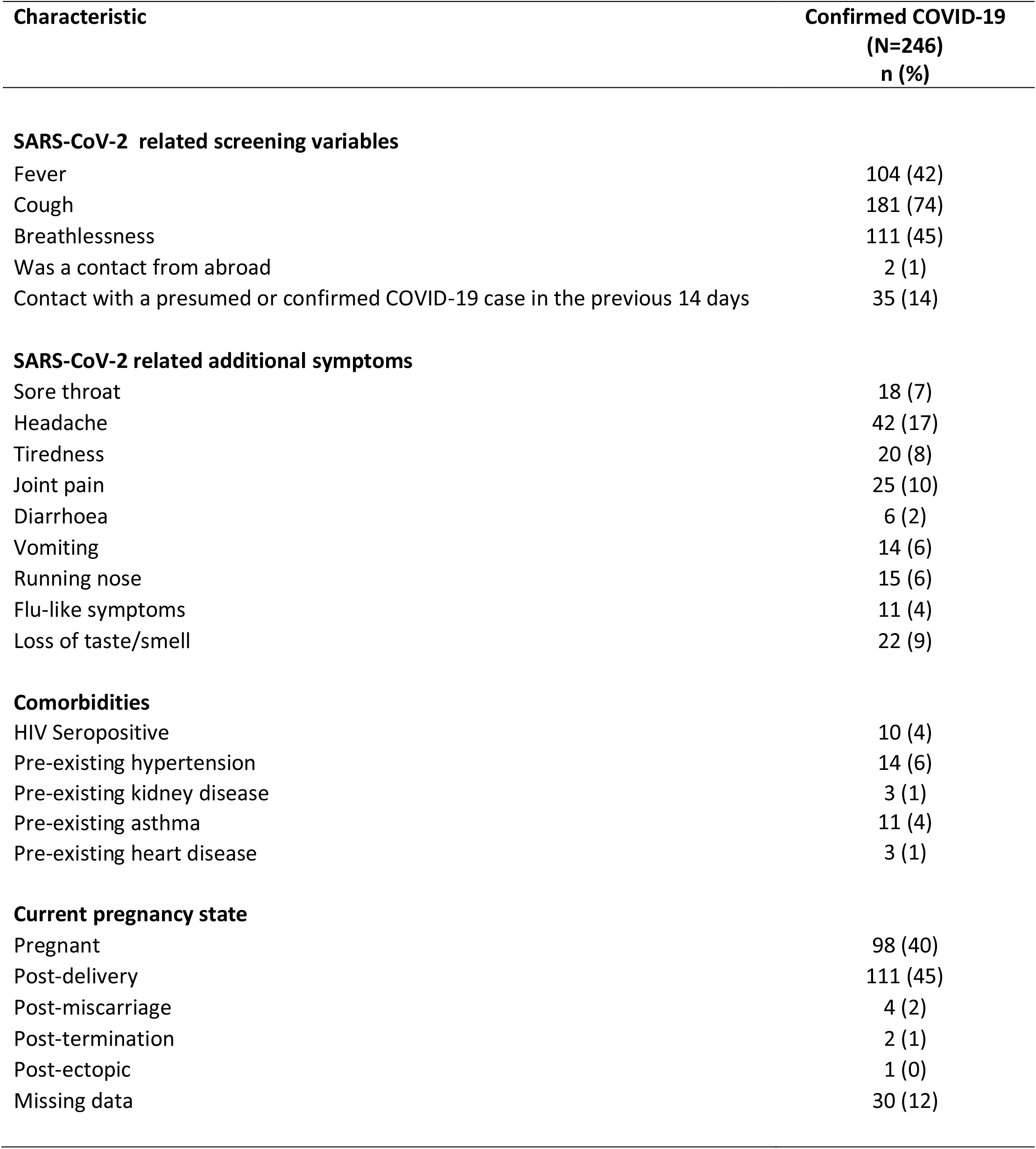
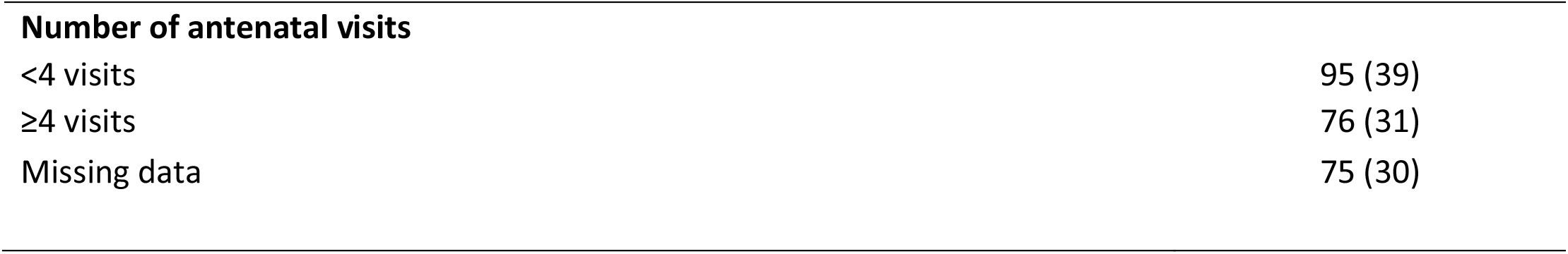
Clinical history and examination characteristics of pregnant and recently pregnant women with SARS-CoV-2 in Malawi.

| Characteristic | Confirmed COVID-19<br>(N=246)<br>n (%) |
| --- | --- |
| **SARS-CoV-2 related screening variables** |  |
| Fever | 104 (42) |
| Cough | 181 (74) |
| Breathlessness | 111 (45) |
| Was a contact from abroad | 2 (1) |
| Contact with a presumed or confirmed COVID-19 case in the previous 14 days | 35 (14) |
| **SARS-CoV-2 related additional symptoms** |  |
| Sore throat | 18 (7) |
| Headache | 42 (17) |
| Tiredness | 20 (8) |
| Joint pain | 25 (10) |
| Diarrhoea | 6 (2) |
| Vomiting | 14 (6) |
| Running nose | 15 (6) |
| Flu-like symptoms | 11 (4) |
| Loss of taste/smell | 22 (9) |
| **Comorbidities** |  |
| HIV Seropositive | 10 (4) |
| Pre-existing hypertension | 14 (6) |
| Pre-existing kidney disease | 3 (1) |
| Pre-existing asthma | 11 (4) |
| Pre-existing heart disease | 3 (1) |
| **Current pregnancy state** |  |
| Pregnant | 98 (40) |
| Post-delivery | 111 (45) |
| Post-miscarriage | 4 (2) |
| Post-termination | 2 (1) |
| Post-ectopic | 1 (0) |
| Missing data | 30 (12) |
| **Number of antenatal visits** |  |
| <4 visits | 95 (39) |
| ≥4 visits | 76 (31) |
| Missing data | 75 (30) |

Women were screened for COVID-19, using the following characteristics cough, breathlessness, fever, history of being in contact with a person with COVID-19. Cough, breathlessness and fever were the most frequent screening symptoms observed at 74% (181/246), 45% (111/246) and 42% (104/246), respectively. Headache, joint pain and tiredness were the most reported COVID-19 related additional symptoms at 17% (42/246), 10% (25/246) and 8% (20/246), respectively. Loss of taste/loss of smell, flu-like symptoms and sore throat were reported in some of the women at 9.% (22/246), 4% (11/246) and 7% (18/246), respectively. Existing medical conditions (co-morbidities) were uncommon; only 4% (11/246) and 6% (14/246) of the women had pre-existing asthma and hypertension, respectively. HIV seropositivity was at 4% (10/246) for the women with COVID-19.

The majority (111/246, 45%) of women were in the post-natal period, and of those who were currently pregnant, 31% (76/246) had attended more than four antenatal visits.

### Clinical, Maternal and Neonatal Outcomes in pregnant and recently pregnant women with COVID-19 infection

Clinical, maternal and neonatal outcomes findings are shown in Table 3. 31% (76/246) of all the women with COVID-19 had a severe maternal outcome. 12% (29/246) suffered a near-miss complication and 19% (n=47/246) died. 6% (14/246) of the women had an oxygen saturation of < 90% for more than one hour, 3% (8/246) had a respiratory rate > 40 or <6 breaths/minute, and 2% (4/246) received mechanical ventilation, not due to anaesthesia. Over half (53%, 130/246) had SARs-CoV-2 virus, related symptoms as their primary reason for hospitalisation.

**Table 3:** Clinical, maternal and neonatal outcomes in pregnant and recently pregnant women with SARS-CoV-2 in Malawi.

| **Outcomes** | **Confirmed COVID-19 (N=246)**<br>**n (%)** |
| --- | --- |
| **Maternal outcomes** |  |
| Severe maternal outcomes | 76 (31) |
| Maternal death | 47 (19) |
| Maternal near-miss complication | 29 (12) |
| No adverse event | 170 (69) |
| **COVID-19 related near-miss lung outcomes** |  |
| Cyanosis | 3 (1) |
| Gasping | 1 (0) |
| Respiratory rate > 40 or < 6 breaths/ minute | 8 (3) |
| O2 saturation < 90 for more than one hour | 14 (6) |
| Requires ventilation not due to anaesthesia | 4 (2) |
| **SARs-CoV-2 related symptoms as primary reason for hospitalization** | 130 (53) |
| **Other reasons for admission** | 61 (25) |
| **Alternative diagnosis\*** | 58 (24) |
| **Number of births (n=111)** |  |
| Singleton | 106 (95) |
| Twins | 4 (4) |
| Triplets | 1 (1) |
| **Birth outcome (n=111)** |  |
| Live births | 101 (90) |
| Macerated still births | 5 (5) |
| Fresh still births | 4 (4) |
| Induced abortion | 1 (1) |
| **Neonatal deaths<sup>†</sup>** | 12(12) |
| Preterm/low birth weight | 0 (0) |
| Asphyxia | 4 (4) |
| Infection | 1 (1) |
| Congenital defects | 1 (1) |
| Other reasons | 1 (1) |
| Unknown | 5 (5) |

Birth outcome and neonatal death is reported for the first birth outcome in an event of multiple gestation
\*Alternative diagnosis means alternative diagnosis to COVID-19 disease that fully explains the clinical presentation
† Number of live births (n=101) is the denominator for the Neonatal deaths

Perinatal outcomes of the 111 births were recorded for the women with COVID-19 infection. From the 111 birth outcomes recorded, 21/111 women (19%) had an adverse perinatal outcome. These consisted of 9/111 (8%) of births being stillbirths; and 12/101 (12%) of the live births resulting in neonatal deaths. Of the neonatal deaths (n=12), the majority were attributed to birth asphyxia and unknown causes at 33% (4/12) and 42% (5/12), respectively.

## DISCUSSION

We report the findings of the Malawian National Maternal Surveillance platform, and its characterisation of the clinical manifestation and outcomes of all pregnant and recently pregnant women hospitalised with confirmed COVID-19 in Malawi between 2nd June 2020 and 1st December 2021 across all district and central hospitals in Malawi.

Notably, 31% (76/246) women with confirmed COVID-19 in this cohort had a severe maternal outcome, of which 19% (47/246) died, and 12% (29/246) suffered a near-miss event. 19% (21/111) of women who delivered during their episode of admission had adverse perinatal outcomes, including 8% (9/111) of women who delivered having a stillbirth and 12% (12/101) of babies dying before discharge from hospital.

The findings in this cohort are stark evidence on the differing impacts and huge disparities of COVID-19 related maternal and neonatal outcomes across the world. Outcomes for COVID-19 positive women are far worse in this cohort than those expected in global data. A population-based cohort of 673 South African (Middle income Country) women admitted to hospital with COVID-19, reported that 32/39 (82%) of deaths were due to COVID-19 infection (5). The perinatal outcomes, 3% (5/148) stillbirths, 3% (5/143) neonatal deaths, from this South African study were also much lower than what we have reported. In addition, it is notable that even in high income settings, Black women had worse outcomes following infection (16).

The Malawian National Maternal Surveillance platform only reported women with COVID-19 who were hospitalised. This enabled the efficient and reliable reporting at scale of those women who were suffering from the most severe disease, but clearly only represents a small proportion of all those women infected. The stigma associated with COVID-19 and challenges in accessing and affording care means it is likely that only those with the most severe symptoms, or who also required hospital care for other pregnancy related reasons were included in this cohort (17). A phone survey of 4641 women and men from 27 districts across all three regions of Malawi, reported that in Malawi, 81% felt they would be treated poorly if they contracted COVID-19 (17). The very high rates of adverse outcomes must also be understood in the context of a maternity care system that even outside of the pandemic was under severe human and physical resource constraints, with maternal mortality rates at 349 per 100,000 births (18), well short of the required Sustainable development goal target of 70 deaths per 100,000 live births (19). In addition to adverse outcomes being directly caused by COVID-19, we also report women for whom COVID-19 was not the primary cause for admission, but nevertheless this diagnosis could have further complicated their care and contribute to the overall high rates of severe outcomes. For example, it could have delayed access to required surgical care or limited access to intensive care capacity as patients with confirmed COVID-19 were cared for in separate clinical areas for infection prevention and control purposes.

Though Malawi prioritised response to the pandemic, the wider impacts throughout the healthcare system were still profound with many preventative and community services disrupted, reduced access to care, as well as staff shortages due to sickness and isolation after being in contact with an individual with COVID-19 (20). In Malawi, there was no widespread community testing for asymptomatic pregnant or recently pregnant women, nor is it possible to disaggregate the national testing data to separately identify those women who were pregnant or recently pregnant. Therefore, the overall levels of asymptomatic or less severe disease in pregnant and recently pregnant women from the wider population during this time period is not known. It was also notable in our cohort that most women admitted to hospitals with COVID-19 did not have known comorbidities. This of course reflects the younger age of this population, and also widespread under ascertainment of chronic disease. Nevertheless, it underlines that health messages on vaccination and other COVID-19 infection prevention measures should target the whole population and that women without known co-existing diseases should not be complacent.

To the best of our knowledge, this is the first national facility-based cohort study of the impact of COVID-19 on pregnant women in SSA. It comprehensively describes the maternal characteristics and clinical outcomes in pregnant and recently pregnant women using data systematically and prospectively collected. It is also important evidence that national surveillance and real-time data provision for government and decision-makers from pregnant women is feasible in a low resource country and can be rapidly established. The success of the implementation of this approach at scale and at speed was due to the commitment of the Ministry of Health, its collaboration with technical partners and the integration with the existing well established Maternal Death Surveillance and Response network (21).

This was also efficient as the data was collected in every facility by the existing “safe motherhood coordinator” who already had similar responsibilities as part of their role for maternal death reporting and reviews. We sought to optimise the quality of data collection by providing in-person training alongside follow-up mentorship and real-time monitoring and feedback of data timeliness and quality. However, as the data relied on existing staff taking on these additional duties it is susceptible to underreporting. This may have been particularly problematic at times of increased service strain such as during the COVID waves and the extent of this effect cannot be readily quantified. This initiative leaves an important legacy in that maternal adverse outcomes beyond those caused by COVID-19 are now electronically recorded and the data rapidly accessible.

During the course of the data collection there was the introduction of lateral flow testing at many of the facilities, which was an important adjunct given the restricted access to molecular testing. These rapid tests were sometimes used at the bedside, but also sometimes used within the hospital laboratory. The type of COVID test undertaken may therefore not have been reliably reported by staff, and lateral flow test results were not universally confirmed with molecular testing, which might have led to misclassification of cases. This paper does not report vaccination status of the women because COVID-19 vaccines were only rolled out in March 2021 and recommended for pregnant women since August 2021 in Malawi. However, this information is now being recorded and will be reported in future. The MATSurvey platform required the daily review of patient whilst they remained in hospital, however, there was no provision for following up patient after discharge, so important outcomes such as neonatal outcomes were only available for those women who delivered during that hospital admission.

In conclusion, this paper addresses a critical gap in the global and regional COVID-19 data, and its impact on the important group of pregnant and recently pregnant woman. The study ascertains that maternal mortality and perinatal outcomes are alarmingly high in a cohort of pregnant and recently pregnant women admitted to Malawian healthcare facilities; a low income country in Africa. In addition, it demonstrates that national surveillance and real-time data provision from pregnant women is feasible in a low resource country and can be rapidly and successfully established.

## Supporting information

Appendix I : Screenshot of the MATSurvey (MATSURV) dashboard

## Data Availability

Due to confidentiality and sensitivity, the raw data used for this study cannot be publicly shared. This is within the mandate of the Research Ethics Committee approval. Requests to access data can be made by contacting the Malawi Ministry of Health and the Malawi Liverpool Wellcome Clinical Research Program.

## AUTHOR CONTRIBUTIONS

All authors contributed to this study’s conceptualisation, writing, and editing and approved the final version for publication. Conceptualisation: CC, LMn and DL. 1^st^ draft writing: CC, LMn and DL. Editing and Review: All authors. Final Script: CC, LMn and DL. Data Curation: CC, DP and LMn. Data analysis: CC and MYRH. Funding acquisition: DL and LG. Supervision: DL. Project management: LG, FK, AL, RB, MM, BM, CN, RM, AK, LM, DP, CM and MK.

## ACKNOWLEDGEMENTS

We thank the Ministry of Health teams at the national, zonal and district levels who ensured the operationalisation of MATSurvey. We especially thank the team of safe motherhood coordinators in the different facilities who tirelessly supported data collection.

## DISCLAIMER

The ideas expressed in this article are of those authors. The funder played no role in the planning or writing of this manuscript.

## ETHICAL APPROVAL

Data gathered through the platform was fully anonymised and made available to the research team by permission of the Ministry of Health, Malawi and the College of Medicine Research Ethics committee (COMREC, P.11/20/3186).

## PROVENANCE AND PEER REVIEW

Externally peer-reviewed.

