## Supplementary figures and images for "The impact of COVID-19 on pregnant and recently pregnant women in Malawi: A national facility-based cohort"

### Appendix I : Screenshot of the MATSurvey (MATSURV) dashboard

**Appendix I : Screenshot of the MATSurvey (MATSURV) dashboard**

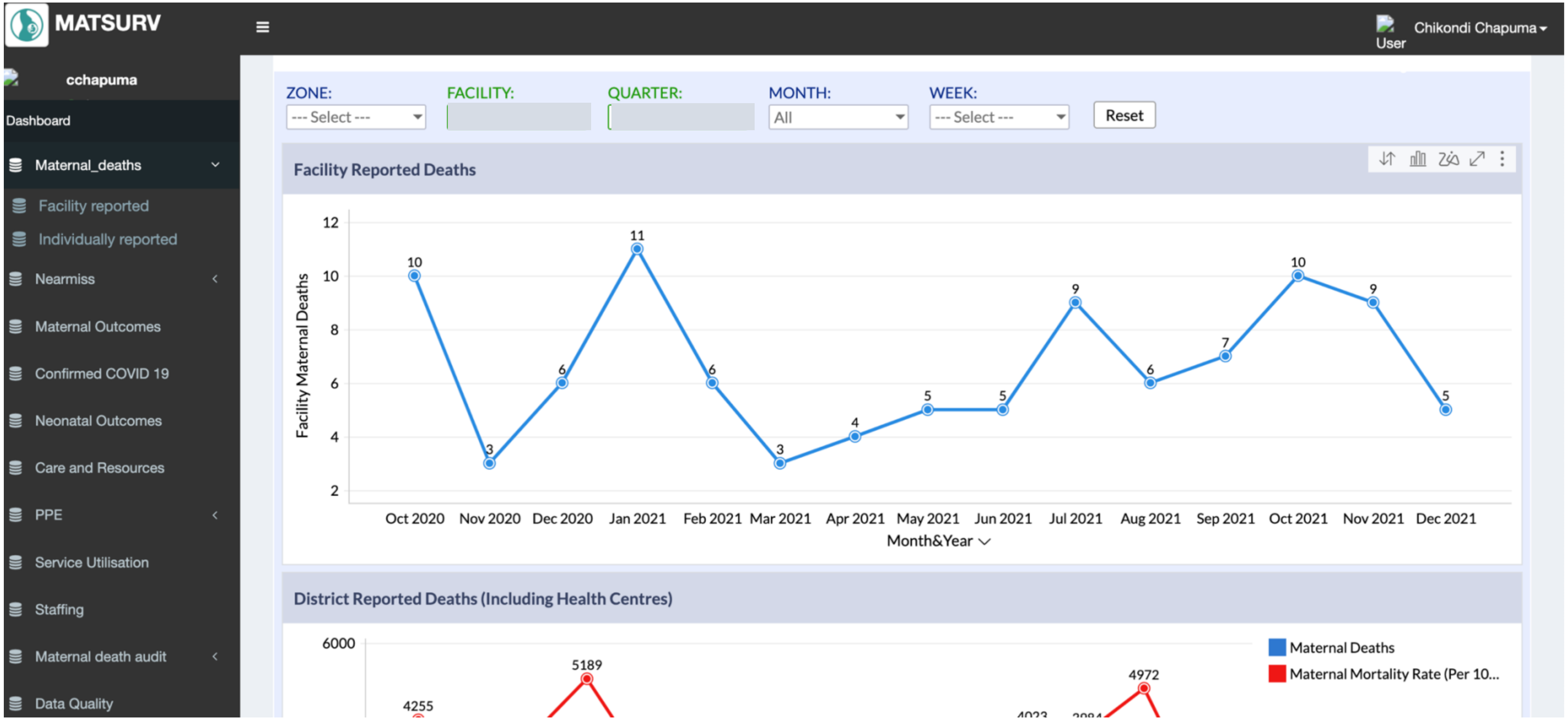
